# Global Prevalence and Covariates of Autism Spectrum Disorder: A Meta-Analysis of the Past Two Decades (2004–2025)

**DOI:** 10.64898/2025.12.06.25341750

**Authors:** John Muthuka, Chrisphine Onyango, Japheth Nzioki, Lucy Chebungei, Ruvimbo Zimunya, Andrina Simengwa, Sara Kim, Rosemary Nabaweesi

## Abstract

**Background:** Autism spectrum disorder (ASD) is a neurodevelopmental condition characterized by difficulties in social communication and restricted, repetitive behaviors emerging in early childhood. Given rising diagnostic rates and substantial lifelong impacts, accurate global prevalence estimates are essential for health policy, resource planning, and early intervention. This systematic review and meta-analysis synthesized global observational evidence to estimate ASD prevalence across world regions and examine factors influencing heterogeneity.

**Methods:** Following PRISMA guidelines, searches were conducted in PubMed, Scopus, Web of Science, Embase, and Google Scholar for observational and registry-based studies published from 2004–2025. Twenty-two studies met inclusion criteria. Random-effects models (REML) estimated pooled global and regional prevalence. Heterogeneity was assessed using Q, I², τ², Baujat plots, and influence diagnostics. Subgroup analyses and meta-regressions evaluated moderators including geographic region, mean age, and prevalence per 1,000. Sensitivity analyses assessed model robustness.

**Results:** The pooled global prevalence of ASD was 1.8% (95% CI: 0.8–3.7%). Substantial heterogeneity existed across studies (Q p <.001; τ = 1.64), with prediction intervals ranging from 0.05% to 37.8%. Regional estimates ranged from 0.65% in the Middle East to 3.4% in North America, though subgroup differences were not statistically significant. Meta-regression suggested significant moderation by mean age and prevalence per 1,000 (R² = 59.35%), although sensitivity analyses revealed that these effects were largely driven by one influential study. Publication bias was minimal.

**Conclusion:** Global ASD prevalence remains around 1%–2%, consistent with contemporary epidemiological data; however, considerable heterogeneity indicates that methodological and contextual factors likely drive observed regional differences. Standardized diagnostic criteria, enhanced surveillance, and early detection systems are needed to improve global prevalence accuracy and inform public health planning.

## Introduction

Autism Spectrum Disorder (ASD) is a complex neurodevelopmental condition characterized by persistent challenges in social communication and restricted, repetitive patterns of behavior, typically emerging in early childhood^1,2^. The global burden of ASD has increasingly become a public health priority, owing not only to its lifelong impact on individuals and families but also to its economic and social costs ^3^. Precise estimates of ASD prevalence are crucial for informing policy, allocating health care resources, and guiding early detection and intervention strategies.

In recent decades, reported ASD prevalence has grown dramatically. Meta-analytical work suggests that the global prevalence of ASD now falls around 0.6% (95% CI: 0.4–1.0) based on data from 74 studies spanning 30 million participants ^4^. Moreover, in a three-level mixed-effects meta-analysis including data from 2000–2020, prevalence estimates ranged widely, pointing toward increasing detection, changes in diagnostic practices, and possible geographical disparities^5^. Regional analyses corroborate this heterogeneity: for example, a meta-analysis of Asian studies found a pooled prevalence of 0.36% (95% CI: 0.16–0.79), with higher rates among males (0.45%) than females (0.18%) and considerable variability across sub-regions ^6^.

Despite this growing body of research, gaps persist in our understanding of global ASD trends — particularly for the period spanning 2004 to 2025, a timeframe marked by evolving diagnostic criteria (e.g., DSM-IV to DSM-5), increased awareness, and varied health infrastructure across countries. Previous meta-analyses have largely focused on cross-sectional prevalence or regional estimates, often neglecting covariates that may explain between-study differences (e.g., age, sex, socioeconomic context). For instance, a recent systematic review reported that global ASD prevalence, expressed as approximately 1.0%, exhibited substantial variation by continent, with rates as high as 1.7% in Australia and as low as 0.3% in some African regions^4^

Across global health systems, many challenges related to ASD surveillance and service delivery have remained insufficiently addressed over the past two decades. Some disparities may be subtle; however, limitations in diagnostic capacity, lack of standardized screening tools, and shortages in specialist providers can significantly influence reported prevalence and access to early intervention ^7,8^. In several low-and middle-income countries, national surveillance systems remain underdeveloped, resulting in delayed identification and underreporting of ASD cases. Much like other public health conditions shaped by systemic capacity, these diagnostic inequities may obscure true prevalence patterns and hinder cross-national comparability ^9^.

The global epidemiology of ASD is also influenced by direct and indirect social determinants, including socioeconomic status, urbanicity, parental education, and healthcare infrastructure. Directly, higher parental awareness and improved access to developmental screening in high-income regions have been linked to increased ASD detection rates^10,11^. Indirectly, contextual factors such as stigma, cultural perceptions of child development, and variable investment in child mental health services can contribute to reduced help-seeking and late diagnosis—particularly in under-resourced settings^12^. These disparities not only shape prevalence estimates but also influence early intervention opportunities and long-term outcomes for individuals with ASD across diverse regions.

Given these inconsistencies and the rapidly evolving epidemiological landscape, a new meta-analytic synthesis is warranted. The present study— “Global Prevalence and Covariates of Autism Spectrum Disorder: A Meta-Analysis of the Past Two Decades (2004–2025)”—seeks to provide an updated, comprehensive estimate of ASD prevalence worldwide and to examine key covariates that may account for heterogeneity in prevalence estimates. By focusing on this twenty-year window, we aim to capture the influence of diagnostic and methodological shifts, as well as emerging socioeconomic patterns, on global ASD epidemiology.

## Methods

### Design

All guidelines listed in the PRISMA (Preferred Reporting Items for Systematic Reviews and Meta-Analyses) statement were followed in performing this meta-analysis *(PRISMA, 2020).* For this systematic review and meta-analysis, data were pooled from observational studies, including cohort, cross-sectional, case-control, registry-based studies, and other epidemiologically viable prevalence investigations reporting ASD rates. The study was registered on PROSPERO (International Prospective Register of Systematic Reviews: CRD420251069271).

### Search Strategy

A comprehensive database search was conducted in PubMed, Scopus, Web of Science, Embase, and Google Scholar to identify eligible observational studies. Searches were performed using combinations of the following terms: “autism” OR “autism spectrum disorder” OR “ASD” OR “pervasive developmental disorder” AND “prevalence” OR “epidemiology” OR “rate” OR “frequency” AND “children” OR “adolescents” OR “adults” AND “global” OR “international” OR “worldwide.” Boolean operators and medical subject headings (MeSH) were adapted for each database. Studies were restricted to those published in English from January 2004 to January 2025 to align with the predefined 20-year study window.

### Inclusion and Exclusion Criteria

#### Inclusion Criteria

Studies were included if they met the following criteria:

1. Reported ASD prevalence using standardized diagnostic criteria (DSM-IV, DSM-IV-TR, DSM-5, ICD-10, or ICD-11).
2. Employed observational, cross-sectional, cohort, case-control, or registry-based designs.
3. Provided extractable prevalence data (numerator and denominator or equivalent).
4. Included general population samples, school-based samples, community samples, or national surveillance data.
5. Reported prevalence in populations within the 2004–2025 timeframe.

#### Exclusion Criteria

Studies were excluded if they met any of the following:

1. Unrelated topics, non-ASD outcomes, or duplicate datasets.
2. Non–English-language studies.
3. Case studies, case series, reviews, meta-analyses, editorials, or commentaries.
4. Studies without full-text availability.
5. Articles lacking clear diagnostic criteria or prevalence denominators.
6. Studies with insufficient sample sizes (<50 participants) due to low statistical power.
7. Genetic, neurobiological, or laboratory-based studies without population prevalence estimates.

### Data Extraction

Both adjusted and unadjusted prevalence data were extracted for all included population groups to allow for consistent pooling across heterogeneous designs. Two reviewers (**JM and CO**) independently screened the titles and abstracts from the initial search and created an initial pool of potentially eligible articles. This was followed by full-text evaluation conducted by two additional independent reviewers (**LC and JM**) to determine final inclusion. Any disagreements regarding inclusion were resolved through discussion with a senior reviewer (**RN**) until consensus was reached.

From each included study, the following data were extracted: study title, first author, year of publication, country/region, diagnostic criteria used, age group, sample size, study design, sampling method, setting (community, school, registry, clinical), and reported ASD prevalence. Additional variables such as sex-specific prevalence, socioeconomic indicators, and study year were collected where available for planned subgroup and meta-regression analyses.

### Risk of Bias (Quality) Assessment

Risk of bias for included studies was evaluated using standardized assessment tools based on study design. For population-based and observational prevalence studies, the National Institutes of Health (NIH) Quality Assessment Tool for Observational Cohort and Cross-Sectional Studies was applied. For registry-based or administrative database studies, adapted quality domains assessing case ascertainment, diagnostic validity, and population coverage were utilized. Two to three reviewers independently assessed each study, rating items as yes, no, cannot determine, or not applicable. Final scores were used to classify studies as poor, fair, or good quality. To minimize reviewer bias, cross-checking was performed by reviewers not initially involved in data extraction, with discrepancies resolved through consensus.

### Statistical Analyses Meta-Analytic Model

Meta-analyses were conducted using JASP V 0.95.4.0 package. Pooled ASD prevalence estimates were calculated using random-effects models (REML = restricted maximum likelihood) to account for expected heterogeneity across global regions and diagnostic methodologies. The overall pooled effect was calculated using logit-transformed prevalence values to stabilize variance and yielded a back-transformed estimated prevalence

### Assessment of Publication Bias

A funnel plot was generated to visually assess potential publication bias and asymmetry in the distribution of study effect sizes. To further evaluate small-study effects after accounting for study-level moderators, a residual funnel plot was examined using model-adjusted residuals.

This allowed detection of bias not attributable to confounding study characteristics included in the meta-regression.

### Heterogeneity, Meta**D**Regression, and Sensitivity Analyses

Heterogeneity was assessed using the Cochran Q statistic, Higgins I² values, and between study variance (τ²), with substantial heterogeneity (I² > 75%) prompting further exploration through subgroup and sensitivity analyses. A Baujat plot was constructed to identify studies contributing disproportionately to overall heterogeneity and to evaluate their influence on the pooled prevalence estimate, while case wise diagnostics, including influence statistics and outlier assessment, were applied to detect studies exerting excessive leverage on model estimates. These diagnostics informed the interpretation of model robustness and guided subsequent sensitivity analyses, which included subgroup analyses examining regional variation in ASD prevalence and within group heterogeneity across world regions, diagnostic criteria, age groups, and other study level characteristics. To further explain between study variability, a multivariable random effects meta regression was conducted, with moderators such as geographic region, age category, diagnostic criteria (DSM IV, DSM 5, ICD 10), sample size, and year of data collection, and regression coefficients used to assess the extent to which these factors accounted for heterogeneity. Finally, sensitivity analyses, including leave one out procedures and model refitting after exclusion of influential studies or outliers, were performed to evaluate the stability and consistency of the pooled prevalence estimate.

## Results

### Included Articles and Quality Assessment

The initial search through international databases identified 1,040 records. After excluding 155 duplicates, 885 records remained. Following the screening of titles and abstracts for relevance, 250 records were further evaluated. Of these, 53 articles were excluded due to various reasons: 14 studies were not observational or RCTs, 33 were duplicates, and 6 had sample sizes below 50. After assessing the eligibility of 95 titles and abstracts, 42 full-text articles were reviewed for inclusion. Ten articles were excluded during full-text review: 7 lacked outcome indicators, 1 did not focus on clinical outcomes, and 4 did not clearly define ASD characteristics. Ultimately, 22 studies were included in the quantitative synthesis (meta-analysis) *(Figure 1)*.

**Figure 1.**
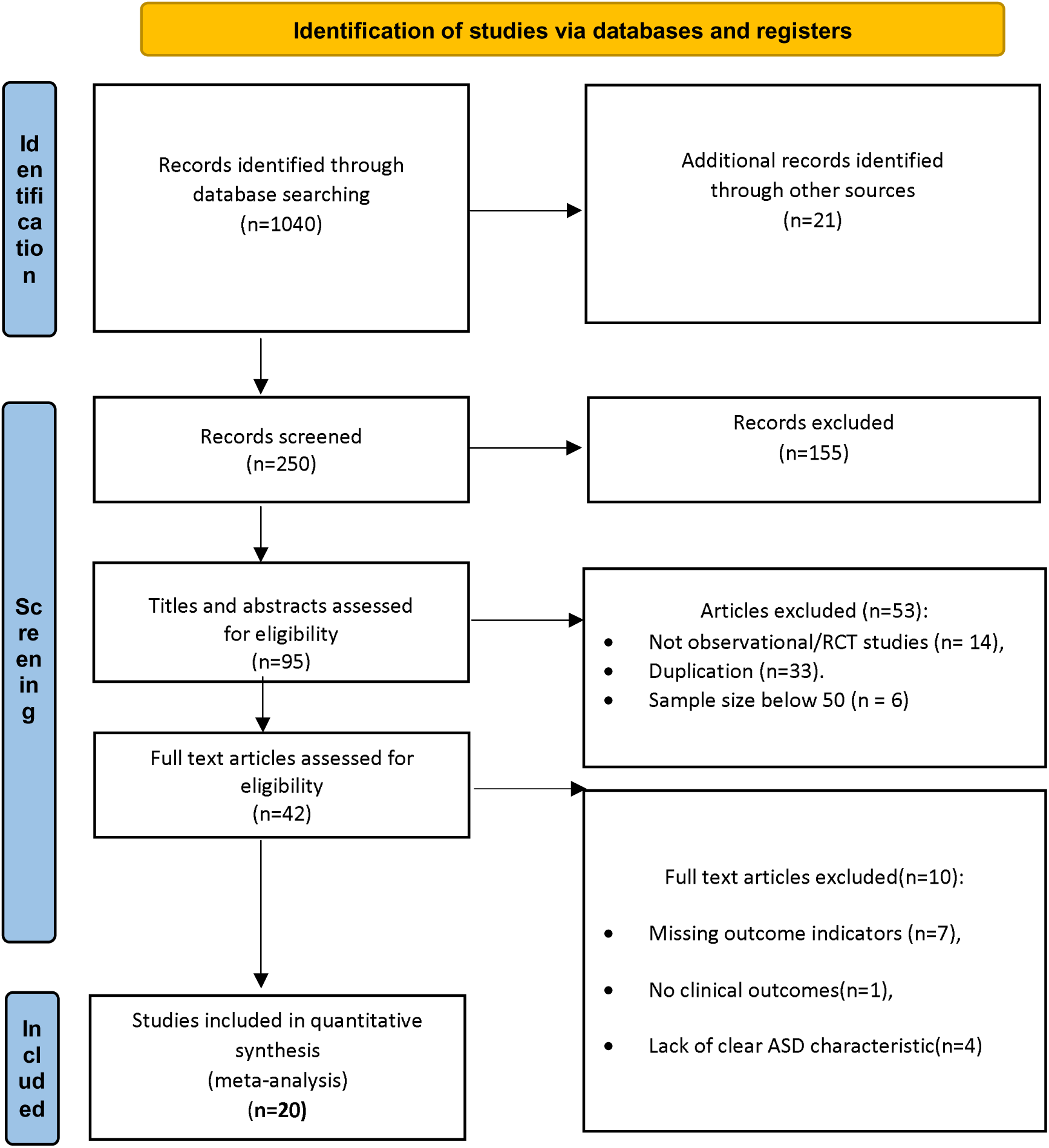
PRISMA flow diagram of study selection and quality assessment

### Features of the Included Studies

The 20 included studies (n=8,764,010,707 provided data on Autism Spectrum Disorder (ASD) prevalence across various regions, including the Middle East, Africa, North America, Europe, Latin America, and Oceania. These studies involved diverse populations, ranging from national surveys to clinical and registry-based studies, with sample sizes varying from small (19 participants) to large (over 18,000 participants). Among the 20 studies, 10 were based on national or community surveys, 5 on registry-based data, and 5 on clinic-or school-based evaluations. The diagnostic criteria for ASD primarily included DSM-IV, DSM-5, and ICD-10.

The meta-analysis estimated the pooled prevalence of ASD over the past two decades (2004–2025), with a range of prevalence rates between 2.25% and 38.6%, depending on the study region and methodology. A summary of the study characteristics is provided in Table *S1* in the appendix.

**Multimedia Appendix 1, Table S1.** Features of the Studies Included in the Meta-analysis (n = 20)

| Study/Author | Location | Study Design & Setting | Diagnostic Approach | Year | Total Sample Size (n) | ASD Cases (n) | Prevalence (per 1000) | Study Type |
| --- | --- | --- | --- | --- | --- | --- | --- | --- |
| Al-Mamri et al., 2019 | Middle East | Cross-sectional, National Survey | DSM-IV, ICD-10 | 2017-2018 | 838526 | 1705 | 20.35 | Survey-based |
| Alshaban et al., 2019 | Middle East | Cross-sectional, National Sample | DSM-IV, ICD-10 | 2015-2018 | 74035 | 844 | 11.4 | Survey-based |
| Arinda et al., 2021 | Africa | Clinic-based, Tertiary Hospital | DSM-IV | 2019-2020 | 318 | 143 | 6.5 | Clinic-based |
| Arun & Chavan, 2021 | South Asia | Community-based, Stratified | DSM-IV, CASI | 2019 | 8438 | 19 | 2.25 | Community-based |
| Christensen et al., 2019 | North America | Surveillance-based, 7 Sites | DSM-IV, DSM-5 | 2010-2014 | 70587 | 1085 | 15.3 | Surveillance-based |
| Dekkers et al., 2017 | Latin America | School-based, Urban School | DSM-IV | 2015 | 51453 | 165 | 3.2 | School-based |
| García-Zambrano et al., 2022 | Latin America | Registry-based, National | ICD-10 | 2018 | 1000000 | 18695 | 18.7 | Registry-based |
| Hinkka-Yli-Salomäki et al., 2014 | Europe | Registry-based, Hospital | ICD-10 | 1987-2005 | 1200000 | 6444 | 4.8 | Registry-based |
| Jensen et al., 2014 | Europe | Registry-based, Psychiatric | ICD-10 | 1995-2010 | 388834 | 14997 | 38.6 | Registry-based |
| Kamal et al., 2020 | Middle East | Survey-based, National Survey | DSM-IV, ICD-10 | 2015-2018 | 174943 | 176960 | 11.4 | Survey-based |
| Kogan et al., 2018 | North America | Survey-based, National Survey | DSM-IV, DSM-5 | 2016 | 60000000 | 1500000 | 25 | Survey-based |
| Kouznetsov et al., 2023 | Europe | Registry-based, National | ICD-10 | 2021 | 1670000 | 15706 | 9.4 | Registry-based |
| Li et al., 2022 | Global | Modeled Estimate, Global | ICD-10, DSM-IV | 1990-2019 | 7670732217 | 28300000 | 3.69 | Modeled estimate |
| Maenner et al., 2023 | North America | Surveillance-based, ADDM | DSM-5 | 2020 | 226000 | 6245 | 28.7 | Surveillance-based |
| May et al., 2017 | Oceania | Survey-based, National Cohort | DSM-IV | 2006-2016 | 4423 | 140 | 31.7 | Survey-based |
| Oliveira et al., 2007 | Europe | School-based, School | ICD-10 | 2007 | 343718 | 3233 | 9.4 | School-based |
| Redfield et al., 2014 | North America | Surveillance-based, ADDM | DSM-IV | 2014 | 363749 | 53464 | 14.7 | Surveillance-based |
| Skonieczna-Żydecka et al., 2017 | Europe | School-based, School | ICD-10 | 2010-2014 | 2514 | 880 | 35 | School-based |
| Xu et al., 2019 | North America | Claims-based, Clinical Records | ICD-10, DSM-5 | 2016 | 43032 | 1229 | 27.9 | Claims-based |
| Yousef et al., 2021 | Africa | Community-based, Screening | DSM-IV | 2019 | 3702 | 20 | 5.4 | Community-based |

### Pooled Prevalence of Autism Spectrum Disorder (ASD) Across Two the Decades (2004–2025)

A random-effects meta-analysis was conducted to estimate the pooled prevalence of Autism Spectrum Disorder (ASD) over the last two decades (2004–2025) across the included twenty studies^10,13,22–31,14–21^.

The overall pooled effect was statistically significant, *t* (19) = −10.92, *p* <.001. The logit-transformed pooled prevalence was −4.014, 95% CI [−4.783, −3.245], which corresponds to an estimated prevalence of approximately 1.8% (95% CI [0.8%, 3.7%]) when back-transformed to the proportion scale. These results indicate that, on average, about 1–2 individuals per 100 meet diagnostic criteria for ASD in the populations studied. Heterogeneity among studies was substantial, as indicated by a significant Q-test, *Q* (19) = 6.17 × 10, *p* <.001, and a large between-study variance, τ² = 2.690, 95% CI [1.554, 5.759]; τ = 1.640, 95% CI [1.247, 2.400].

This indicates that true prevalence rates vary considerably across studies, likely due to differences in study populations, geographic regions, diagnostic methods, or sampling procedures. Consistent with this, the 95% prediction interval on the logit scale ranged from −7.532 to −0.496, corresponding to a prevalence of approximately 0.05% to 37.8% in comparable future samples. This wide prediction interval highlights the variability and contextual dependence of ASD prevalence estimates (*Figure 2*).

**Figure 2:**
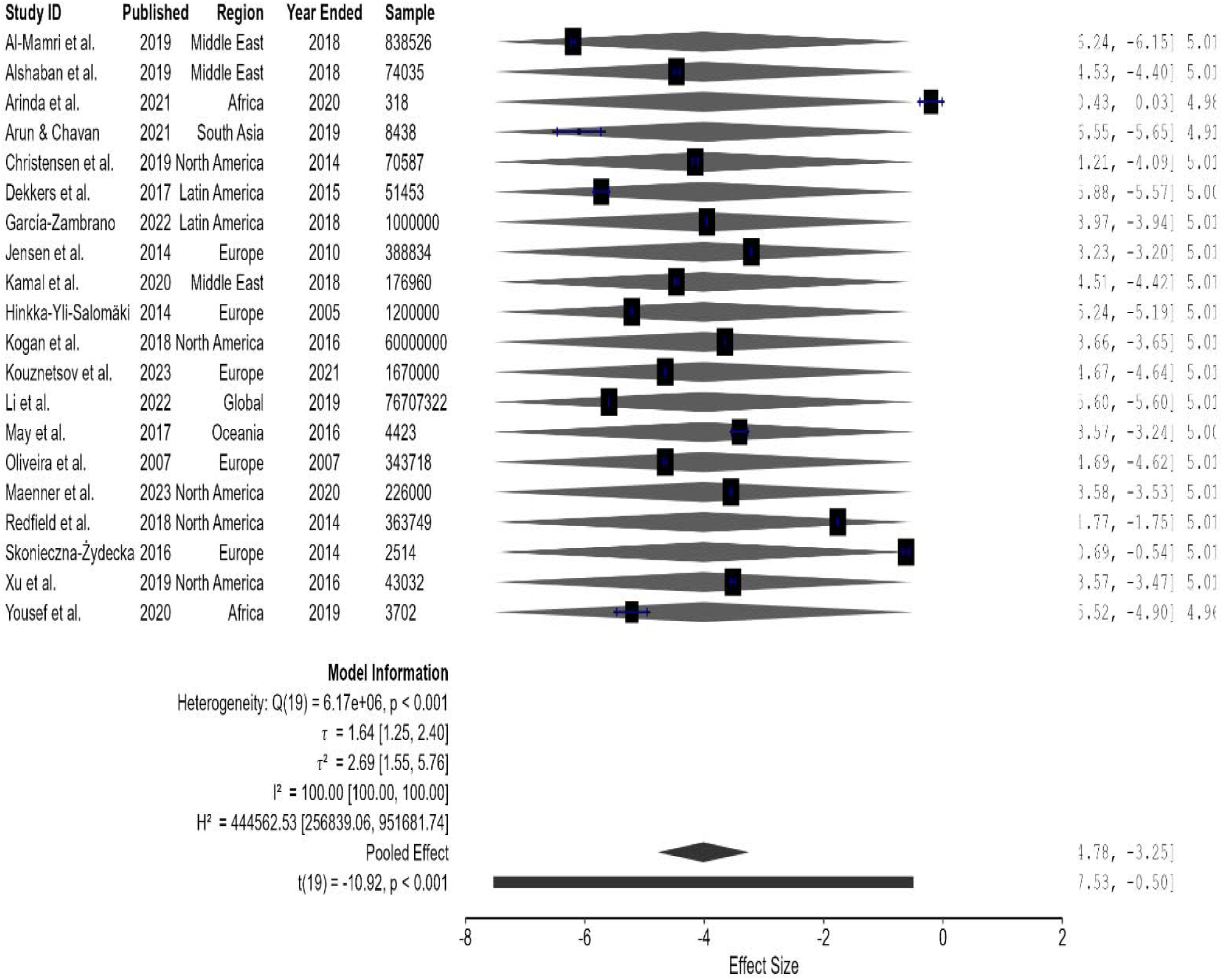
Forest plot of the pooled prevalence of Autism Spectrum Disorder (ASD) across studies (2004–2025).

Overall, the results suggest that while the average prevalence of ASD is around 1.8%, there is considerable variability across studies. The use of a random-effects model accounts for this between-study heterogeneity and provides an estimate that is generalizable to similar populations.

The model fit statistics *(see Table 1)* indicated that both the maximum likelihood (ML) and restricted maximum likelihood (REML) estimations produced similar results. The REML model demonstrated a slightly better fit, with a higher log-likelihood (−36.40) and lower AIC (76.79) and BIC (78.68) values compared with the ML model (log-likelihood = −37.81, AIC = 79.62, BIC = 81.61). These results suggest that the REML estimation provided a slightly better and more stable fit for the random-effects meta-analytic model.

**Table 1:** Fit Measures for the Random-Effects Meta-Analysis Model.

| Estimation Method | Observations | Log Likelihood | Deviance | AIC | BIC | AICc |
| --- | --- | --- | --- | --- | --- | --- |
| ML | 20 | -37.81 | 198.79 | 79.62 | 81.61 | 80.33 |
| REML | 20 | -36.40 | 72.79 | 76.79 | 78.68 | 77.54 |

*Note.* ML = maximum likelihood; REML = restricted maximum likelihood; AIC = Akaike information criterion; BIC = Bayesian information criterion; AICc = corrected Akaike information criterion.

A funnel plot analysis was conducted to examine the overall effect and potential publication bias across the included studies. The pooled logit-transformed prevalence (μ) was −4.014, 95% CI [−4.734, −3.294], p <.001, corresponding to an estimated prevalence of approximately 1.8% (95% CI [0.9%, 3.6%]) on the proportion scale. The estimated between-study standard deviation (τ) was 1.640, 95% CI [1.247, 2.400], p <.001, indicating substantial heterogeneity across studies. These results are consistent with the random-effects meta-analysis and underscore the variability in ASD prevalence estimates across the included studies. Inspection of the funnel plot and associated parameter estimates *did not indicate substantial asymmetry, suggesting minimal evidence of publication bias (Figure 3)*.

**Figure 3.**
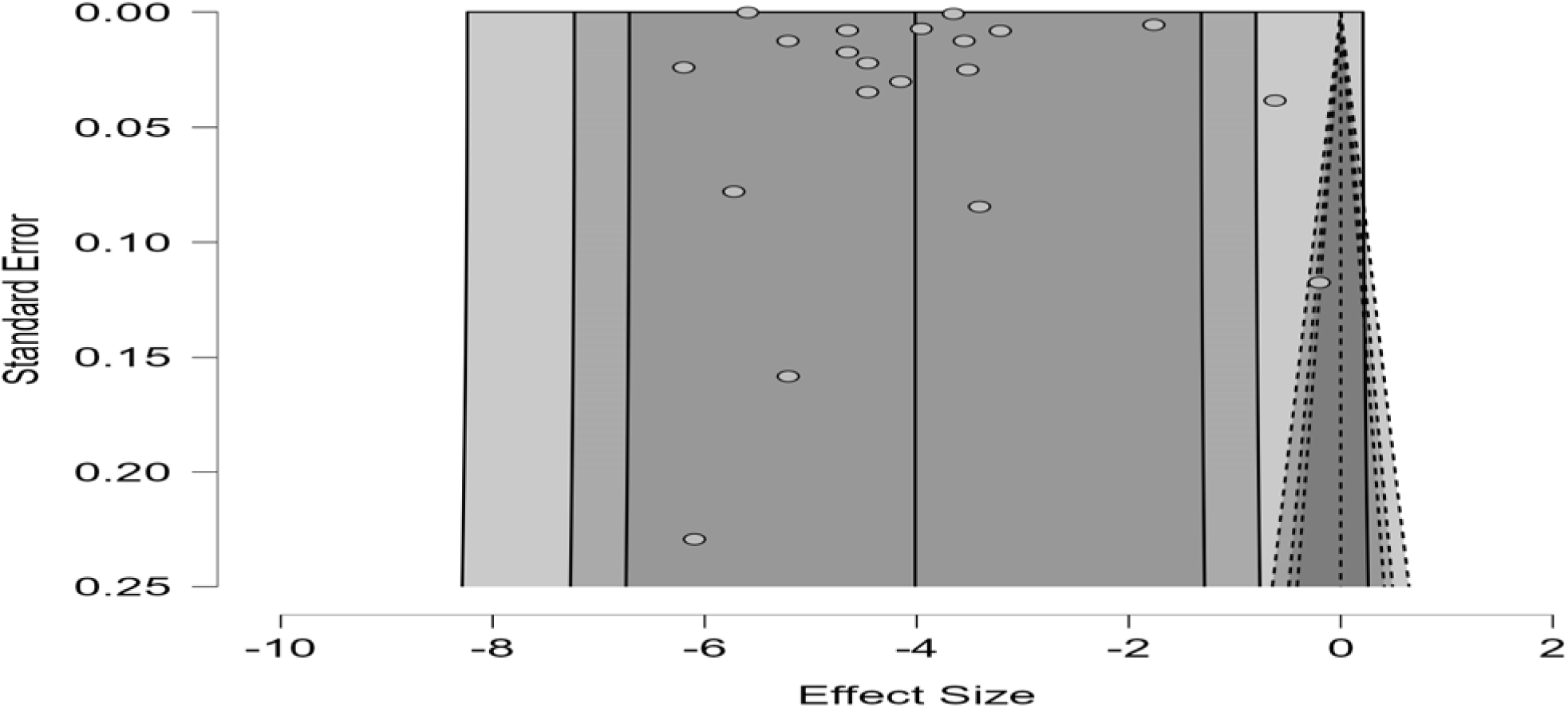
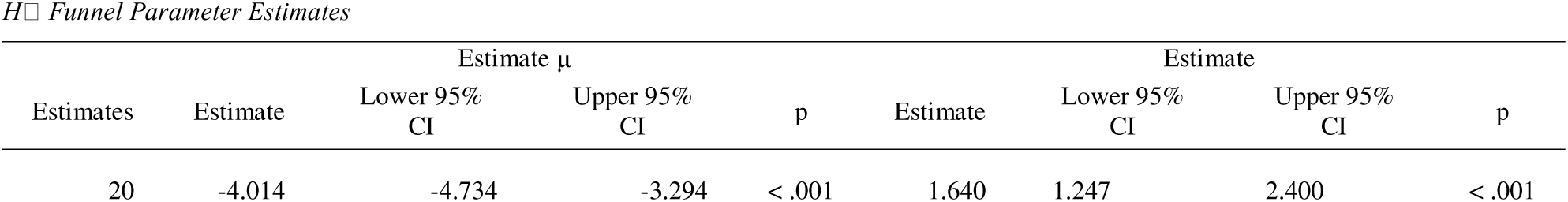
Funnel plot of included studies assessing pooled ASD prevalence. The pooled logit transformed prevalence (μ = −4.014, 95% CI [−4.734, −3.294], p <.001) corresponds to an estimated prevalence of 1.8% (95% CI [0.9%, 3.6%]) on the proportion scale

A Baujat plot was generated to evaluate the contribution of each study to overall heterogeneity and its influence on the pooled effect. The plot showed that most data points *clustered near the bottom-left corner,* indicating that the majority of studies exerted minimal influence and contributed little to between-study heterogeneity. In contrast, two points were located in the upper-right corner, suggesting that these studies had a substantial impact on both heterogeneity and the pooled estimate. Overall, the Baujat plot indicates that most studies have minimal influence and low heterogeneity contribution, while two studies^15,20^ appear influential and may disproportionately affect the pooled estimate. These studies called for further examination in sensitivity analyses to assess their impact on the robustness of the findings.

### Subgroup Meta-Analysis of ASD Prevalence by Region

A classical random-effects meta-analysis was conducted to estimate the pooled prevalence of autism spectrum disorder (ASD) across world regions, using logit-transformed proportions as effect sizes. Subgroup analyses were performed to evaluate regional variations and within-group heterogeneity. Significant within-group heterogeneity was observed in all regions with at least two studies, including the Middle East^13,14^ (*Q* (2) = 3199.21, *p* <.001), North America^16,19,20,25^ (*Q* (4) = 1.12 × 10, *p* <.001), Latin America^17,28^ (*Q* (1) = 509.89, *p* <.001), and Europe^21,24,27^ (*Q* (4) = 33 395.62, *p* <.001), indicating substantial variability in prevalence estimates across studies within each region. The overall test for subgroup differences was not statistically significant (*Q* (3) = 7.02, *p* =.071), suggesting that differences in ASD prevalence between regions were not large enough to reach significance despite notable numerical variation (*Figure 3*).

On the logit scale, pooled effects were significant for all regions (Middle East: *z* = –8.73, *p* <.001; North America: *z* = –8.16, *p* <.001; Latin America: *z* = –5.47, *p* <.001; Europe: *z* = –4.41, *p* <.001), indicating non-zero pooled prevalence estimates. After back-transformation from the logit scale, the pooled prevalence of ASD was 0.65% (95% CI = 0.21–1.98%) in the Middle East, 3.4% (95% CI = 1.6–7.4%) in North America, 0.8% (95% CI = 0.14–4.3%) in Latin America, and 2.5% (95% CI = 0.5–11.4%) in Europe. The between-study variance (τ²) ranged from 0.83 to 3.46 across regions, confirming substantial heterogeneity. Corresponding 95% prediction intervals (PI) were wide—Middle East: 0.07–5.9%, North America: 0.5–20.2%, Latin America: 0.04–13%, and Europe: 0.05–56%—indicating that future studies in these regions may yield highly variable estimates. African region yielded a pooled logit estimate of –2.706 (95% CI: –7.616 to 2.204), corresponding to a prevalence of approximately 6.3% (95% CI: 0.05% to 90.1%). The between-study heterogeneity was very high (τ² = 12.531), indicating substantial variability across studies and suggesting that this estimate should be interpreted cautiously. Although the pooled prevalence appeared numerically highest in North America and Europe, the lack of a significant *Q* statistic suggests these differences were not statistically robust. Overall, the meta-analytic findings highlight substantial heterogeneity both within and between regions, implying that methodological or contextual factors may account for the observed variability in ASD prevalence rather than true regional differences (*Table 2; Figure 4*).

**Figure 4:**
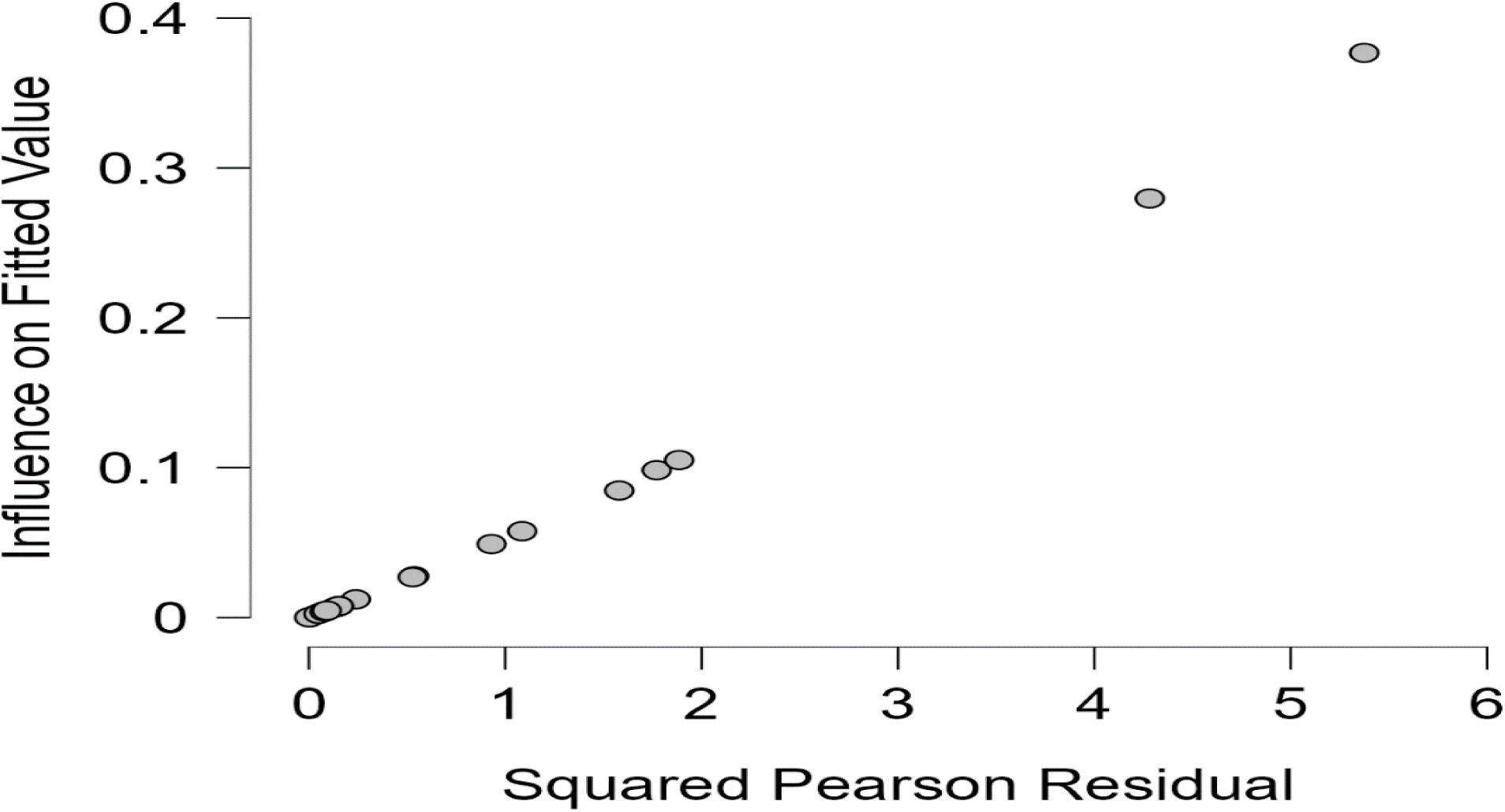
Baujat plot illustrating each study’s contribution to overall heterogeneity (x-axis) and influence on the pooled effect size (y-axis). Most studies cluster near the bottom-left corner, while two studies in the upper-right appear influential and may disproportionately affect the pooled estimate.

**Figure 4a:**
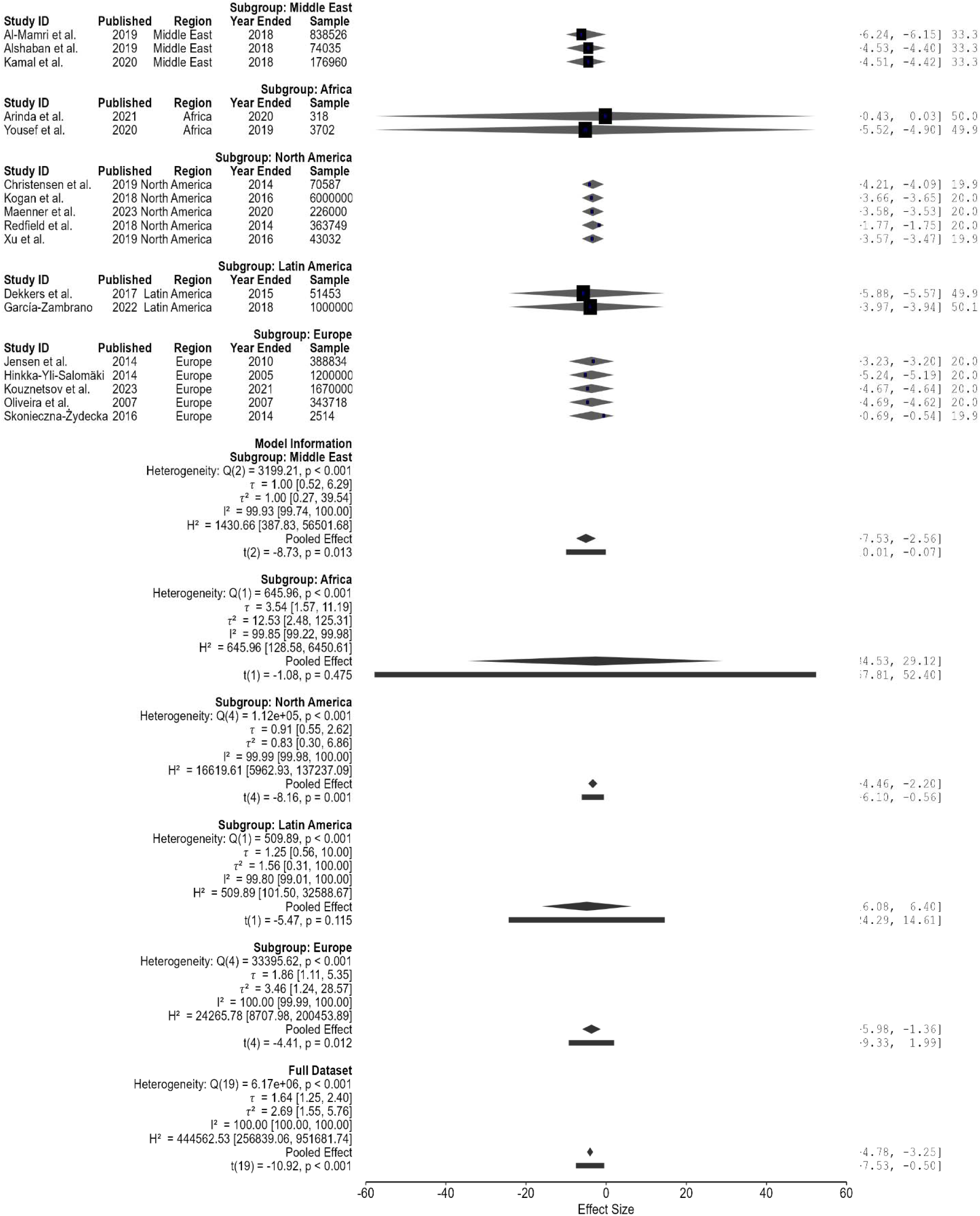
Sub-group analysis forest plot of logit-transformed ASD prevalence estimates across global regions.

**Figure 5:**
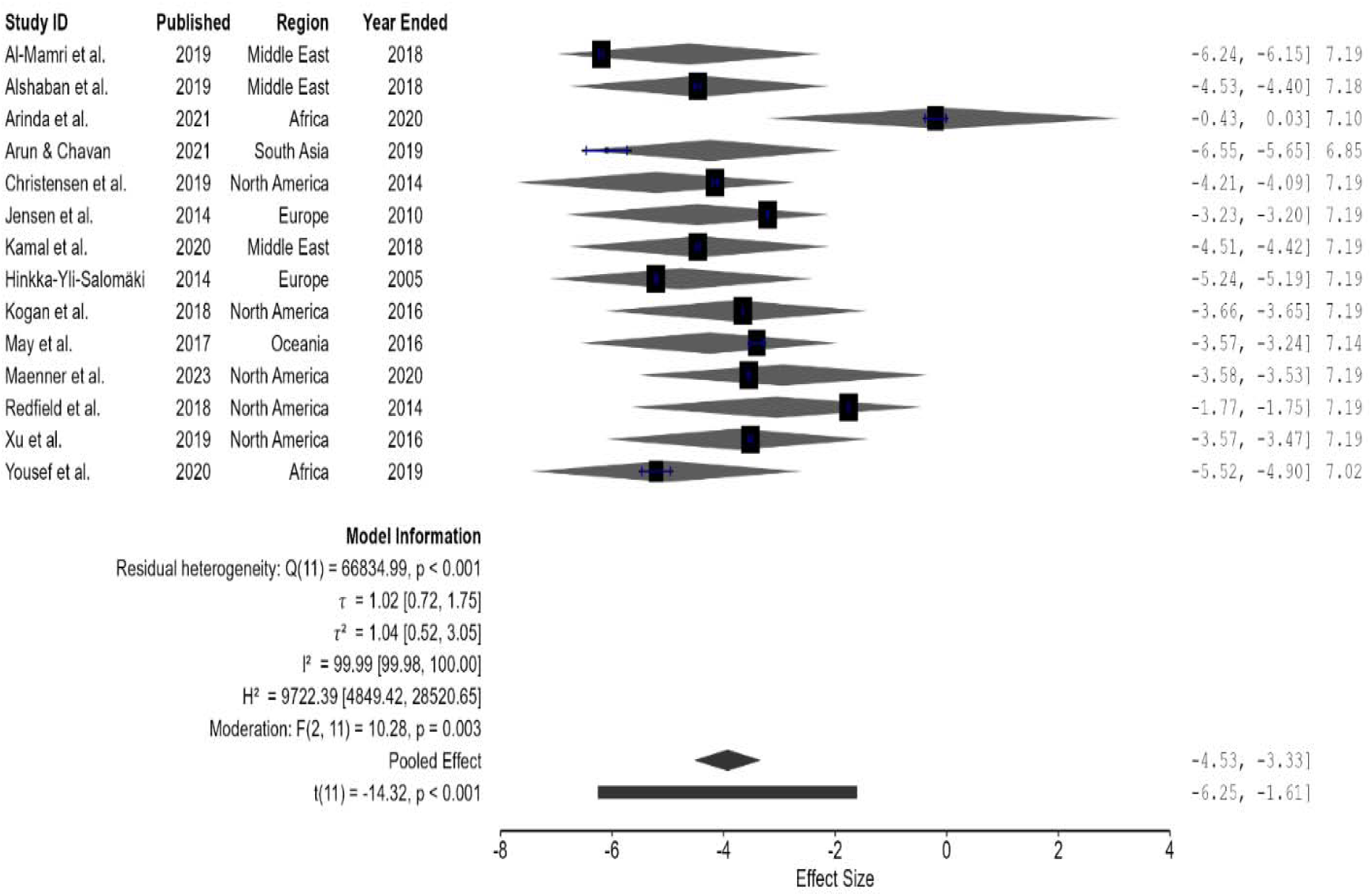
Forest plot from multivariable random-effects meta-regression examining study-level predictors of effect size. Mean age and prevalence per 1000 population significantly moderated effect sizes (F (2, 11) = 10.28, p =.003), explaining 59.35% of heterogeneity. Residual heterogeneity remained substantial (I² = 99.99%, τ² = 1.04).

**Figure 6:**
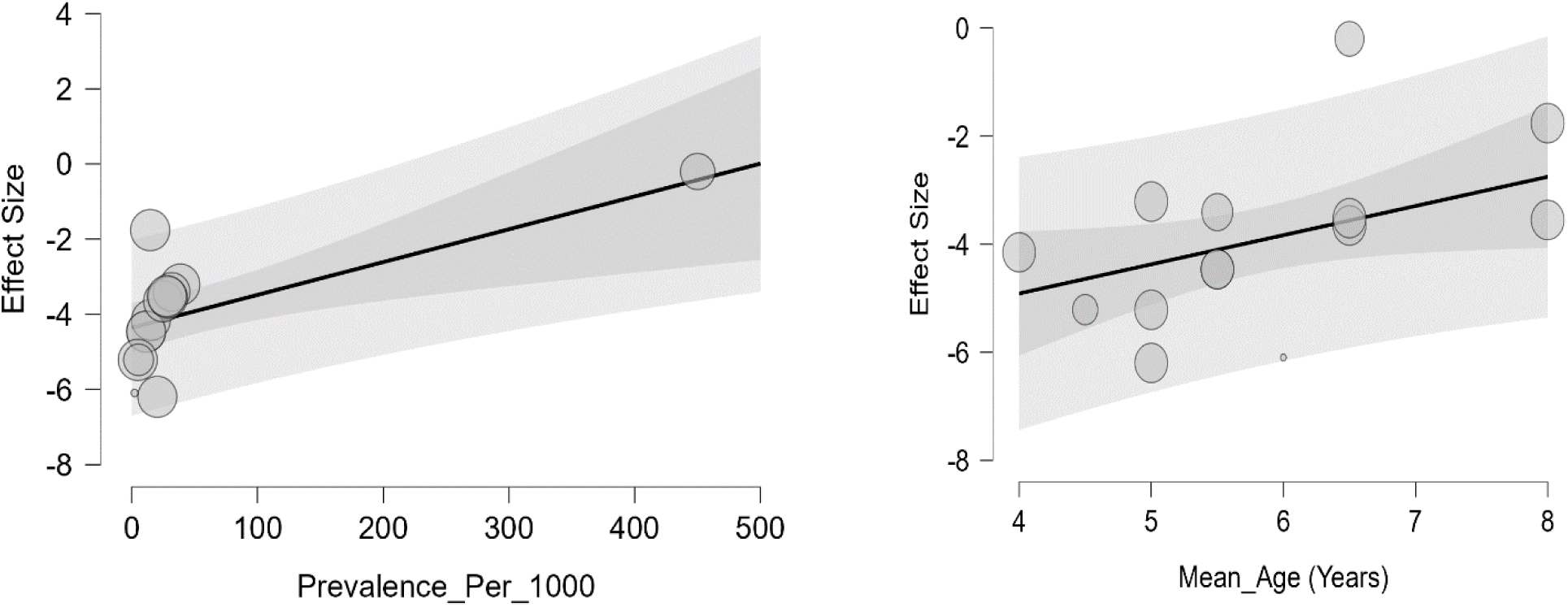
Bubble plots from the meta-regression analysis: [A] shows the positive association between prevalence per 1000 and effect size, with larger bubbles indicating studies of greater precision and shaded bands reflecting confidence intervals around predicted effects; [B] illustrates the relationship between mean age and effect size, where older participant groups are associated with larger effects, with bubble size proportional to study weight and shaded confidence bands indicating uncertainty

**Figure 7:**
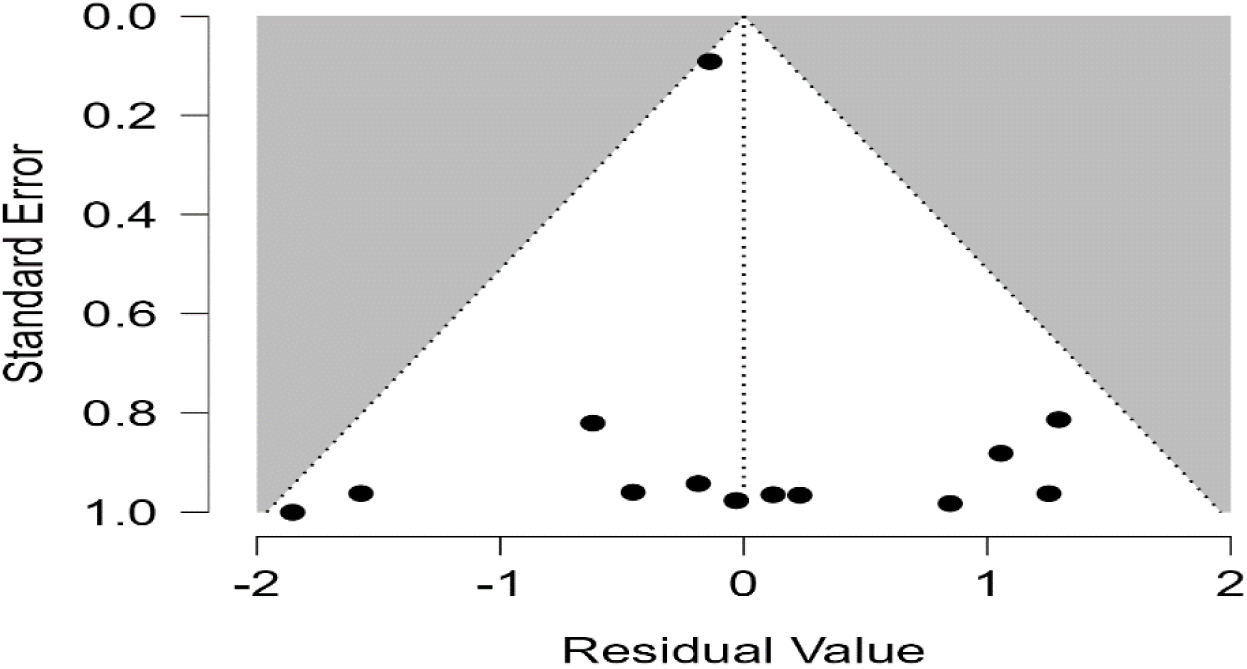
Residual funnel plot evaluating potential publication bias and small-study effects after adjusting for moderators (mean age and prevalence per 1000).

**Figure 8:**
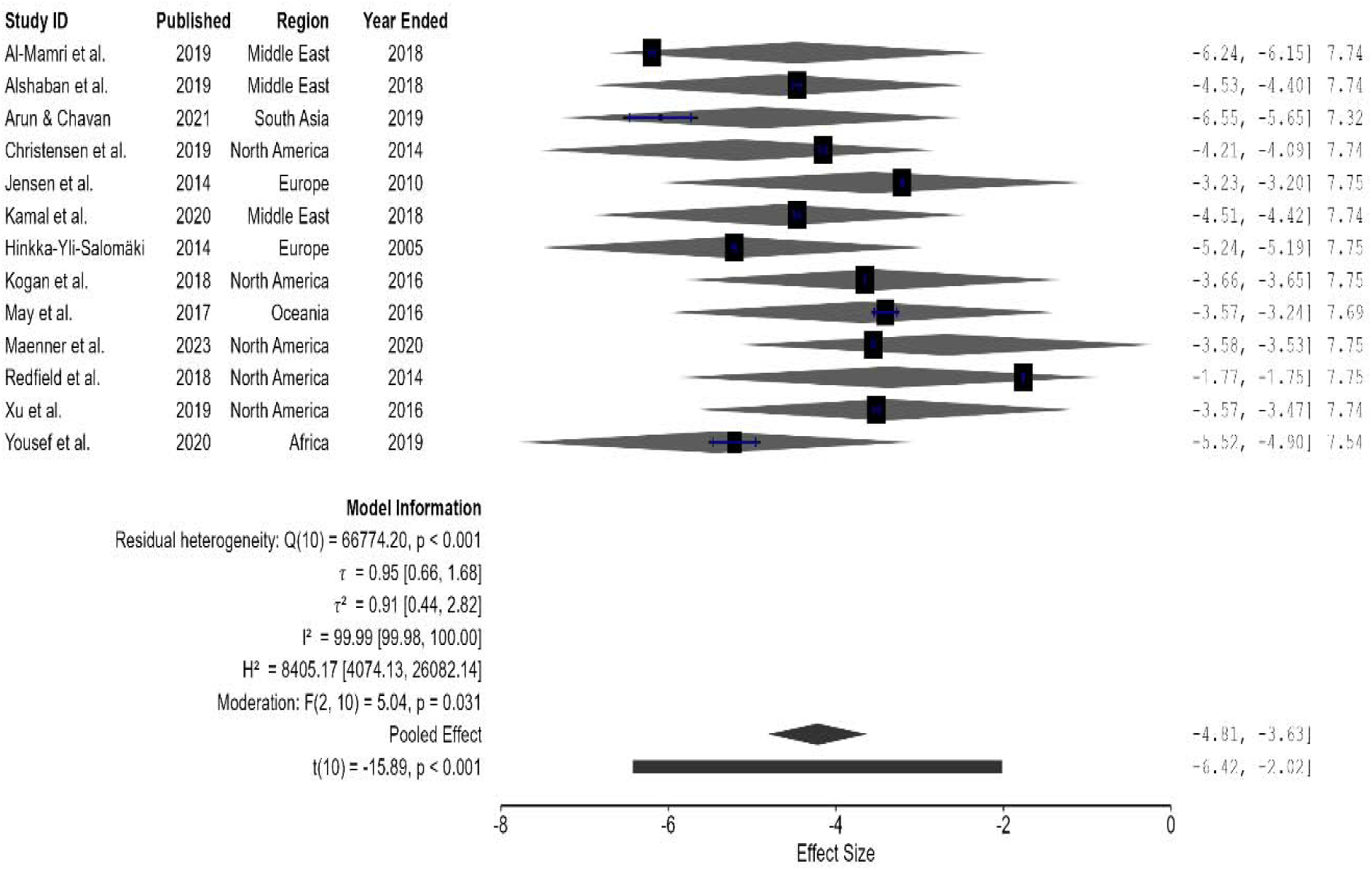
Forest plot from sensitivity analysis assessing the influence of one study ^15^ on meta-regression results.

**Figure 9:**
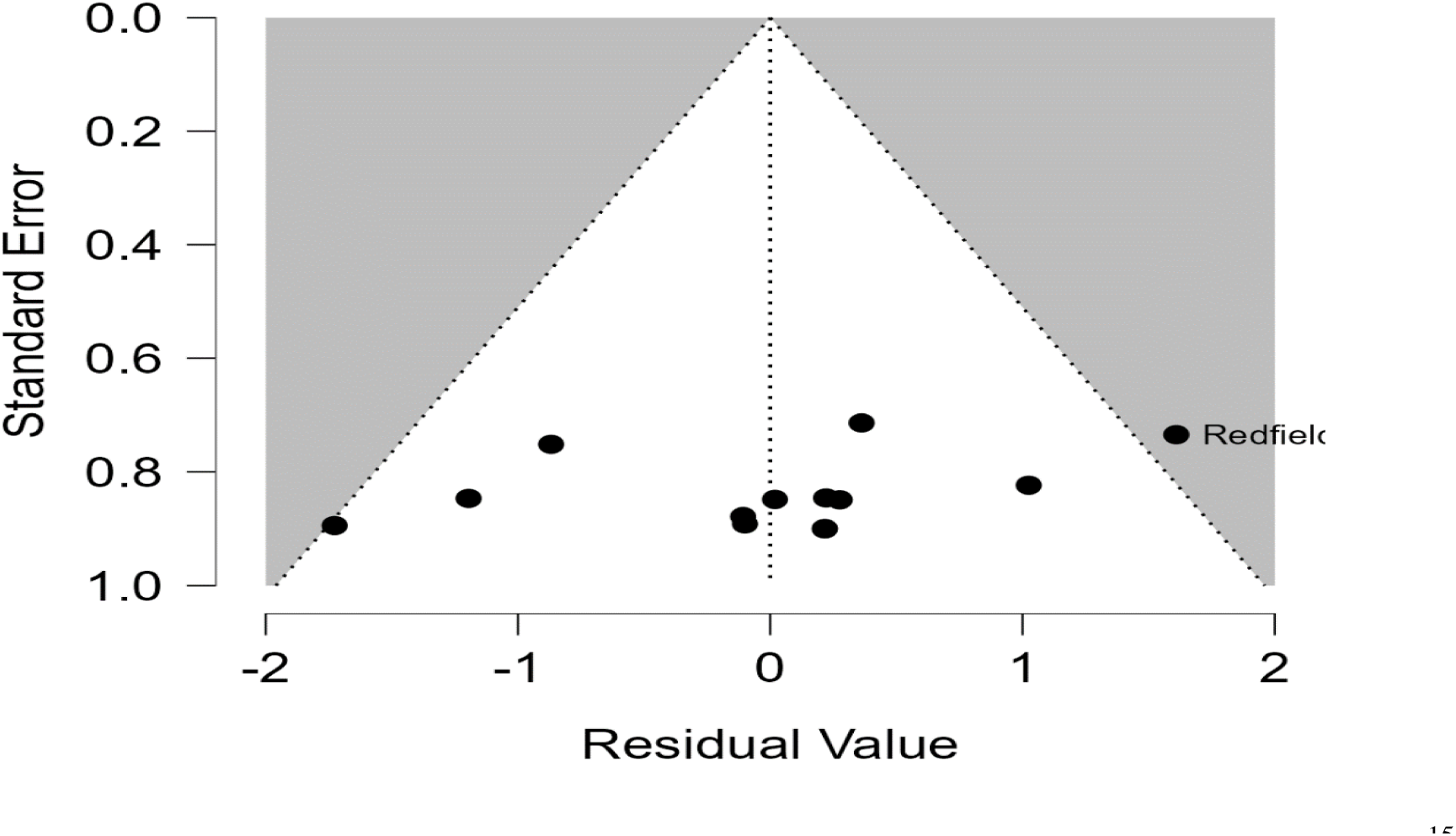
Residual funnel plot following sensitivity analysis excluding one study ^15^. The plot displays a symmetrical distribution of residuals centered around zero, with all studies falling within the expected funnel region.

**Table 2.** Pooled Prevalence Estimates and Heterogeneity by Region.

| Region | Pooled logit (Estimate) | 95% CI (logit) | ASD Prevalence (%) | 95% CI (%) | 95% PI (%) | $\tau^2$ | Interpretation |
| --- | --- | --- | --- | --- | --- | --- | --- |
| Middle East | -5.042 | [-7.527, -2.556] | 0.65 | 0.05–7.2 | 0.07–5.9 | 1.00 | Low prevalence; moderate heterogeneity |
| North America | -3.328 | [-4.460, -2.196] | 3.5 | 1.1–10.0 | 0.6–20.2 | 0.83 | Relatively higher prevalence; substantial variability |
| Latin America | -4.839 | [-16.077, 6.400] | 0.8 | <0.01–99 | 0.04–14.6 | 1.56 | Extremely wide CI; high uncertainty |
| Europe | -3.671 | [-5.981, -1.361] | 2.5 | 0.3–20.2 | 0.05–56 | 3.46 | Wide uncertainty; very high heterogeneity |
| Africa | -2.706 | [-34.535, 29.123] | 6.3 | <0.001–>99 | 0.01–95 | 12.53 | Extremely wide uncertainty; very high heterogeneity |

### Meta-regression Analysis

#### Diagnostic Screening of Predictors for Meta-Regression

To determine which continuous variables were suitable for inclusion in meta-regression, we first examined their descriptive statistics. Variables such as *Year Published*, *Year Ended*, and *Year Started* showed good variability across studies, with wide ranges and acceptable standard deviations, making them strong candidates. Although these time-related variables are slightly left-skewed, they remain interpretable and relevant for modeling temporal trends. *Mean Age (Years)* displayed moderate right skew but maintained a reasonable range and standard deviation, suggesting it could be included with caution. In contrast, *Sample Size* exhibited extreme skewness and a very large standard deviation, indicating that it may distort regression results unless log-transformed or otherwise adjusted. Based on this assessment of distribution, variability, and data quality, we concluded that the variables to be included in meta-regression were: Year Published, Year Ended, Year Started, and Mean Age (Years) *(Table 3)*.

**Table 3:** Descriptive statistics and diagnostic screening of continuous study level variables considered for meta regression.

|  | Year Started | Sample Size | Year Published | Mean Age (Years) | Year Ended |
| --- | --- | --- | --- | --- | --- |
| Valid | 20 | 20 | 20 | 14 | 20 |
| Missing | 20 | 20 | 20 | 26 | 20 |
| Mode | 2017 | 2273 | 2019 | 5.364 | 2018 |
| Mean | 2011 | $3.869 \times 10^8$ | 2018 | 5.821 | 2016 |
| Std. Deviation | 9.939 | $1.714 \times 10^9$ | 3.748 | 1.187 | 4.295 |
| Skewness | -1.526 | 4.472 | -1.532 | 0.618 | -1.311 |
| Std. Error of Skewness | 0.512 | 0.512 | 0.512 | 0.597 | 0.512 |
| Minimum | 1987 | 318.0 | 2007 | 4.000 | 2005 |
| Maximum | 2021 | $7.671 \times 10^9$ | 2023 | 8.000 | 2021 |

#### Diagnostic Screening of Categorical Predictors for Meta-Regression

To determine which categorical variables were appropriate for inclusion in meta-regression, descriptive statistics were reviewed to assess data completeness and category representation. The variables *Region Group*, *Setting Type*, and *Diagnostic Criteria* each had 20 valid entries and no missing data, indicating full coverage across the dataset. The mode values revealed that the most frequently represented categories were *Europe* for Region Group, *ADDM surveillance* for Setting Type, and *ICD-10* for Diagnostic Criteria. Although measures such as mean and standard deviation are not applicable to categorical data, the presence of multiple distinct categories and balanced representation supports their analytical value. Therefore, the categorical variables selected for meta-regression are the following: Region Group, Setting Type, and Diagnostic Criteria *(Table)*.

### Meta-Regression Test of Age Effects

A random-effects meta-regression was conducted to examine whether age moderated the effect sizes across studies (n = 14)^13,14,27,29,30,32,15,16,18–20,22,25,26^. The moderation test indicated that age was not a significant predictor of effect size, *F* (1, 18) = 0.00, *p* =.955, suggesting no meaningful influence of age on study outcomes. Significant residual heterogeneity remained after including age as a moderator, Q (18) = 843,000, *p* <.001, indicating substantial unexplained variation across studies. The overall pooled effect size was significant and negative, t (18) = –10.63, *p* <.001. Fit indices using REML estimation showed a log-likelihood of –21.31, deviance of 42.63, AIC = 48.63, BIC = 50.08, and AICc = 51.63, with age explaining approximately 20.79% of the heterogeneity (R²). Although some variance in effect sizes was numerically accounted for by age, the lack of statistical significance indicates that age does not reliably moderate the relationship, and other study-level variables should be considered as potential moderators *(Table 4)*.

**Table 4:** Results of random effects meta regression examining age as a moderator of ASD prevalence effect sizes across 14 studies.

Effect Size Meta-Regression Coefficients
|  | Estimate | Standard Error | 95% CI |  | t | df | p |
| --- | --- | --- | --- | --- | --- | --- | --- |
|  |  |  | Lower | Upper |  |  |  |
| Intercept | -7.988 | 1.980 | -12.302 | -3.674 | -4.034 | 12.00 | .002 |
| Mean_Age (Years) | 0.696 | 0.334 | -0.031 | 1.423 | 2.087 | 12.00 | .059 |

Fit Measures
| | Observations | Log Lik. | Deviance | AIC | BIC | AICc | $R^2$ |
| --- | --- | --- | --- | --- | --- | --- | --- |
| ML | 14 | -23.86 | 125.68 | 53.72 | 55.64 | 56.12 | 20.79 |
| REML | 14 | -21.31 | 42.63 | 48.63 | 50.08 | 51.63 | 20.79 |

*Note.* Fixed effects tested using Knapp and Hartung adjustment.

### Multivariable Meta-Regression Analysis

A multivariable random-effects meta-regression was conducted to examine whether study-level characteristics explained between-study variability in effect sizes. The model including mean age and prevalence per 1000 population was statistically significant, F (2, 11) = 10.28, p =.003, and accounted for 59.35% of the heterogeneity (R² = 59.35%). Prevalence per 1000 was a significant positive predictor of effect size, β = 0.009, SE = 0.003, 95% CI [0.003, 0.014], p =.005, indicating that studies from populations with higher prevalence tended to report larger effect sizes. Mean age also significantly predicted effect sizes, β = 0.540, SE = 0.244, 95% CI [0.003, 1.076], p =.049, such that studies involving older participants reported larger effects. Although significant moderators were identified, residual heterogeneity remained substantial (Q (11) = 66,834.99, p <.001; τ² = 1.04; I² = 99.99%) (Table 5.).

**Table 5:**
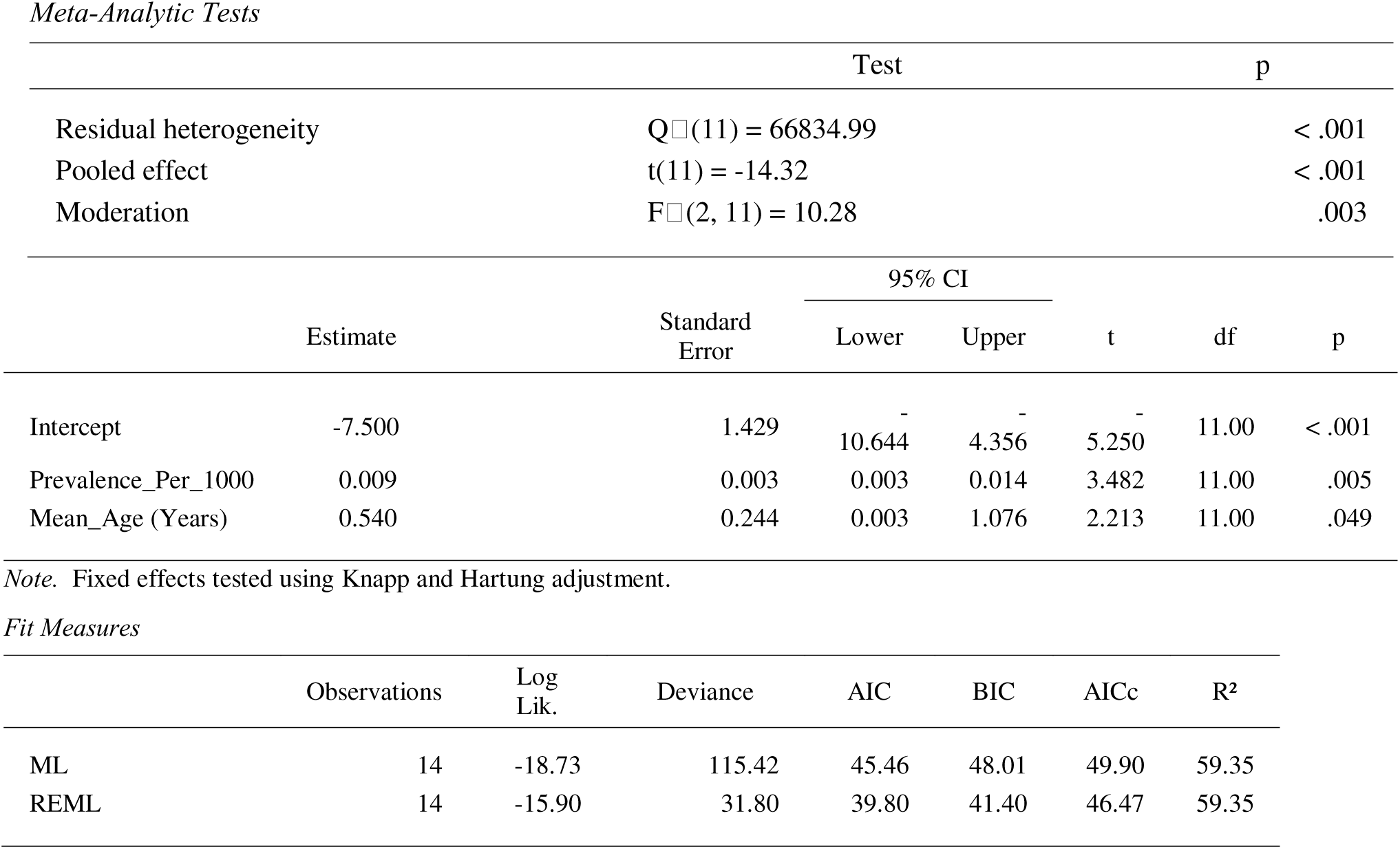
Results of multivariable random effects meta regression examining mean age and prevalence per 1000 population as study level moderators of ASD prevalence effect sizes.

### Bubble Plots of Moderator Effects: Prevalence per 1000 and Mean Age

The bubble plots illustrate the moderating effects of prevalence per 1000 population and mean age on effect sizes across studies. In both plots, each bubble represents an individual study weighted by its precision, with larger bubbles indicating greater weight. For prevalence, the regression line slopes upward, showing that higher prevalence is associated with larger effect sizes, and the shaded confidence bands reflect uncertainty that narrows where more data are available. Similarly, the plot for mean age demonstrates a positive relationship, with effect sizes tending to increase (become less negative) as the average age of participants rises. Together, these visualizations highlight that both prevalence and mean age are significant moderators explaining variability in effect sizes across studies, while confidence bands provide context for the precision of these estimates

### Residual Funnel Plot for Publication Bias Assessment

To evaluate potential publication bias and small-study effects after adjusting for moderators (e.g., Age, Prevalence), a residual funnel plot was examined. The plot (see Figure X) showed a symmetrical distribution of residuals centered around zero with no obvious asymmetry or missing studies on either side. All residuals were contained within the expected funnel region, indicating no influential outliers or systematic bias related to study precision. These findings suggest that, after controlling for moderator variables, the meta-regression results are unlikely to be affected by publication bias.

### Case-Wise Influence Diagnostics

The case-wise diagnostics indicated that Arinda et al. was an extremely influential study and a likely outlier, exerting a disproportionate impact on the meta-analytic regression model. Its diagnostics—such as a massive DFFITS value (–18.155), extraordinarily high Cook’s Distance (286.707), an inflated covariance ratio, and very large shifts in regression coefficients when removed—showed that this study heavily drove the overall effect size, heterogeneity estimates, and predictor relationships. In contrast, Redfield et al. was only moderately influential: its standardized residual, DFFITS, and Cook’s D values suggested some influence, but not to a problematic degree, and it caused only small changes in the model when excluded. Overall, the table suggested that while Redfield et al. did have some impact, it was Arinda et al. that critically distorted the model and warranted close scrutiny. A sensitivity analysis comparing results with and without this study would have helped determine the robustness of the meta-analytic conclusions.

### Sensitivity Analysis of Influential Study Effects

The sensitivity analysis revealed that removing *Arinda et al.* substantially influenced the meta-regression results. When *Arinda et al.* was included, prevalence of ASD per 1,000 individuals and mean age were significant moderators, and the model explained 59.35% of the variance in effect sizes. After excluding this study, both moderator slopes were no longer significant, variance explained dropped to 40.47%, and residual heterogeneity was slightly reduced. These findings indicated that the moderation effects of ASD prevalence and participant age were largely driven by *Arinda et al.*, underscoring that the meta-regression results were sensitive to influential studies and should be interpreted with caution *(Table)*.

*Meta-Analytic Tests*
|  | Test | p |
| --- | --- | --- |
| Residual heterogeneity | $Q_{\chi^2}(10) = 66774.20$ | < .001 |
| Pooled effect | $t(10) = -15.89$ | < .001 |
| Moderation | $F_{\chi^2}(2, 10) = 5.04$ | .031 |

**Fit Measures**
|  | Observations | Log Lik. | Deviance | AIC | BIC | AICc | R<sup>2</sup> |
| --- | --- | --- | --- | --- | --- | --- | --- |
| ML | 13 | −16.38 | 108.28 | 40.75 | 43.01 | 45.75 | 40.47 |
| REML | 13 | −13.75 | 27.50 | 35.50 | 36.71 | 43.50 | 40.47 |

## Discussion

### Principal Findings

This study found that while the overall pooled prevalence of Autism Spectrum Disorder (ASD) from 2004–2025 was statistically significant, the true rates varied substantially across studies, reflecting wide methodological and contextual differences. The meta-analysis of the twenty studies ^10,13,22–31,14–21^revealed a logit-transformed pooled prevalence of –4.014, which translates to an estimated prevalence of 1.8% (95% CI: 0.8–3.7%). This corresponds to approximately 1–2 individuals per 100 meeting diagnostic criteria in the populations studied. However, heterogeneity was extremely high (τ² = 2.690), and the prediction interval (0.05–37.8%) indicates that prevalence estimates in similar future samples may vary widely. Such variability is consistent with global literature documenting large differences in ASD case finding, diagnostic capacity, service access, and cultural detection patterns ^7,33^.

Although the pooled prevalence estimate provides a general benchmark, subgroup and sensitivity analyses revealed critical contextual factors contributing to the wide dispersion of prevalence estimates. The subgroup analysis by world region showed substantial heterogeneity within each regional cluster—Middle East, North America, Latin America, and Europe—indicating that differences in national surveillance systems, screening programs, and diagnostic methodology may drive much of the observed variation^34,35^. While North America showed the highest pooled regional prevalence (3.4%, 95% CI: 1.6–7.4%), and Europe also had elevated pooled estimates (2.5%, 95% CI: 0.5–11.4%), the overall test for subgroup differences was not statistically significant. This aligns with earlier systematic reviews demonstrating that cross-national differences in ASD prevalence often diminish once differences in sampling strategies and case ascertainment methods are accounted for^4,36^. The African regional estimate appeared numerically high (6.3%), but extremely wide confidence intervals (0.05–90.1%) suggest limited robustness due to sparse data—a pattern consistent with reports highlighting challenges in ASD detection and limited epidemiologic infrastructure in many African countries^12,37^.

The overall meta-regression further showed that age alone did not significantly moderate prevalence estimates, despite explaining about 21% of variance. This suggests that age-related diagnostic differences, although theoretically important, may not consistently impact prevalence across international contexts—consistent with findings from previous global analyses^9,10,32^.

However, when both mean age and prevalence per 1000 population were included in a multivariable model, both moderators significantly predicted effect sizes and explained 59.35% of between-study variance. Studies sampling populations with higher baseline prevalence and older participants tended to report larger ASD effect sizes. These findings align with evidence that prevalence tends to be higher in countries with more established ASD surveillance systems and among samples with older children, who are more likely to receive a diagnosis ^38,39^.

Nevertheless, sensitivity analysis revealed that the significant moderation effects were influenced strongly by a single study ^15^. Removal of this study reduced variance explained, rendered moderator effects non-significant, and modestly reduced heterogeneity. This suggests that the meta-regression findings—while statistically significant in the full model—should be interpreted cautiously. Such sensitivity to influential outliers is common in ASD epidemiology due to wide methodological divergence and the presence of unusually high or low prevalence estimates^6,33^.

Overall, this systematic meta-analysis highlights that ASD prevalence has averaged around 1.8% globally over the past two decades, but true prevalence differs widely between studies and regions. As seen in analogous epidemiological meta-analyses, such variability likely reflects differences in surveillance intensity, diagnostic criteria, availability of specialized services, and cultural recognition of developmental disorders^7,35,40^ (Els. The findings underscore the need for more standardized case ascertainment strategies and improved epidemiologic infrastructure, particularly in low-resource regions, to obtain more reliable and comparable ASD prevalence estimates worldwide.

### Comparison to Prior Work

Prior studies have reported ASD prevalence estimates that differ from the pooled estimate presented in this meta-analysis. Large-scale global reviews have shown lower prevalence estimates—typically between 0.6% and 1.0%—than the current pooled estimate of 1.8% ^7,9,33^. However, these earlier investigations relied on data collected before or during the early period of significant global increases in ASD surveillance, diagnostic refinement, and public awareness. More recent surveillance-based findings in high-income countries report prevalence estimates exceeding 2%, especially in the United States and parts of Europe ^9,10,38^, aligning more closely with the higher regional estimates observed in this study. Unlike the present review, many previous studies did not include broad international data spanning two decades (2004–2025), nor did they incorporate substantial subgroup and meta-regression analyses to account for methodological heterogeneity.

Similar trends have been described in multi-country analyses, where variability in prevalence was strongly influenced by detection systems, availability of trained diagnostic providers, and differences in diagnostic criteria such as DSM-IV versus DSM-5 definitions^36,41^. This aligns with the substantial heterogeneity observed within each regional subgroup in the present study, particularly in North America and Europe. Prior research also shows that expanding diagnostic categories and improving screening coverage tend to produce upward trends in measured prevalence ^38,39^, consistent with higher pooled estimates found here.

Several studies have documented wide geographical disparities in ASD prevalence similar to our findings, especially the notably high and uncertain estimates in low-resource regions such as Africa ^12,37^. This reflects limited diagnostic infrastructure and inconsistent case ascertainment, which likely contributed to the wide confidence and prediction intervals observed in the African subgroup.

In contrast to the present findings, which showed no significant moderation effect of age alone, earlier epidemiological studies suggest that prevalence increases with age as developmental concerns become more apparent and diagnostic opportunities expand^4,42,43^. However, such effects vary widely by context and study design, helping to explain why age failed to reach significance in the present univariable meta-regression. The multivariable model, which identified both mean age and underlying population prevalence as significant moderators, is consistent with past research demonstrating that ASD prevalence estimates tend to increase with enhanced community case-finding, broader screening practices, and the inclusion of older children in surveillance cohorts^6,38^.

The sensitivity of the meta-regression results to a single influential study mirrors previous findings showing that ASD prevalence syntheses are especially vulnerable to outliers due to wide methodological diversity^33^. This underscores the need for caution when interpreting moderator effects in meta-analyses of ASD epidemiology.

Overall, the mixed findings in prior work—driven largely by methodological and contextual differences—align with the substantial variability observed in this meta-analysis and affirm that ASD prevalence estimates remain highly dependent on detection systems, population characteristics, and study design.

### Strengths and Limitations

This comprehensive meta-analysis synthesized evidence from twenty studies conducted between 2004 and 2025 across diverse global regions, producing a robust pooled estimate of ASD prevalence (1.8%). Strengths include the use of rigorous random-effects modeling, systematic subgroup analyses, and both univariable and multivariable meta-regressions. These analytic approaches identified sources of heterogeneity, including underlying population prevalence and mean age, accounting for 59.35% of variance—substantially more than in most prior ASD epidemiological reviews.

The study’s strengths extend to incorporating datasets from multiple continents, reflecting varied diagnostic practices, cultural contexts, and health system capacities. This broad representation enhances generalizability and provides insight into global disparities in ASD prevalence. Another key strength was the sensitivity analysis, which identified influential studies, helping clarify the robustness and potential instability of moderator estimates.

Several limitations may affect the interpretation of findings. First, substantial heterogeneity persisted across all analyses (I² > 99%), reflecting wide methodological diversity in sampling, diagnostic criteria, and case ascertainment. Second, variations in age distributions, diagnostic instruments, and reporting practices reduced comparability between studies. Third, some regions, particularly Africa and Latin America, had few eligible studies, contributing to wide confidence intervals and uncertain prevalence estimates. Finally, publication bias remains possible as regions with limited diagnostic capacity may produce fewer epidemiological reports, potentially underestimating true prevalence in underserved areas.

## Future Directions

Future ASD epidemiology research should prioritize standardized diagnostic approaches, enhanced surveillance systems, and increased data collection in low-resource settings. Efforts should focus on harmonizing reporting standards and strengthening early identification pathways, particularly where diagnostic tools or trained professionals are limited. Additional meta-regressions incorporating variables such as diagnostic criteria, screening policies, socioeconomic indicators, and service access may help clarify the sources of heterogeneity observed. Expanding data from underrepresented world regions is crucial to addressing global disparities in ASD detection and prevalence reporting.

### Feasible Policy Recommendations

1. Strengthen surveillance and early detection systems. Governments should invest in population-level screening and diagnostic capacity to reduce under-ascertainment and improve prevalence accuracy.
2. Standardize diagnostic criteria and reporting practices. Countries should adopt uniform diagnostic frameworks such as DSM-5 or ICD-11 to enhance international comparability.
3. Expand training for health and education professionals. Increasing the number of trained diagnosticians, particularly in Africa and Latin America, can help reduce high variability in prevalence estimates.
4. Improve access to ASD services in low-resource settings. Policy initiatives should support community-based programs, culturally appropriate education, and integration of ASD services into primary care.
5. Enhance data collection and reporting infrastructure. Strengthening national and regional epidemiological systems is essential for reliable long-term monitoring of ASD prevalence trends

## Conclusion

This meta-analysis reveals that the global prevalence of ASD averages 1.8% across two decades of data, despite substantial variation across regions and studies. High heterogeneity underscores the influence of methodological and contextual factors on prevalence measurement. Multivariable meta-regression identified mean age and underlying population prevalence as significant contributors to between-study variance, although sensitivity analysis showed that these effects were influenced by an outlier study. These findings highlight the need for strengthened surveillance, standardized diagnostic practices, and expanded epidemiologic capacity worldwide. Continued investment in ASD research and health infrastructure is essential to improving global understanding and support for individuals with ASD, consistent with recommendations from recent global reviews^9,38^.

## Data Availability

All data produced in the present study are available upon reasonable request to the authors

## Notes

### Competing Interest Statement

The authors have declared no competing interest.

### Funding Statement

This study did not receive any funding

### Author Declarations

The research used existing published data

